# CLINICAL PROPERTIES AND DIAGNOSTIC METHODS OF COVID-19 INFECTION IN PREGNANCIES: META-ANALYSIS

**DOI:** 10.1101/2020.06.06.20123901

**Authors:** Banu Uygun-Can, Bilge Acar-Bolat

**Affiliations:** Department of Microbiology, Dental Faculty, Marmara University, Istanbul-Turkey; Department of Quantitative Methods, School of Business, Istanbul University, Istanbul-Turkey

**Keywords:** COVID-19, SARS-CoV-2, pregnancy, clinical characteristics

## Abstract

We aimed to summarize reliable medical evidence by the meta-analysis of all published retrospective studies that examined data based on the detection of severe acute respiratory syndrome coronavirus 2 (SARS-CoV-2) by clinical symptoms, molecular (RT-PCR) diagnosis and characteristic CT imaging features in pregnant women. MEDLINE PubMed, SCOPUS, ISI Web of Science, Clinical Key, and CINAHL databases were used to select the studies. Then, 384 articles were received, including the studies until 01/MAY/2020. As a result of the full-text evaluation, 12 retrospective articles covering all the data related were selected. A total of 181 pregnant cases with SARS-CoV-2 infections were included in the meta-analysis within the scope of these articles. According to the results, the incidence of fever was 38.1% (95% CI: 14.2–65%), and cough was 22% (95% CI: 10.8–35.2%) among all clinical features of pregnant cases with SARS-CoV-2 infection. So, fever and cough are the most common symptoms in pregnant cases with SARS-CoV- infection, and 91.8% (95% CI: 76.7–99.9%) of RT-PCR results are positive. Moreover, abnormal CT incidence is 97.9% (95% CI: 94.2–99.9%) positive. No case was death. However, as this virus spreads globally, it should not be overlooked that the incidence will increase in pregnant women and may be in the risky group. RT-PCR and CT can be used together in an accurate and safe diagnosis. In conclusion, these findings will provide important guidance for current studies regarding the clinical features and correct detection of SARS-CoV-2 infection in pregnant women, as well as whether it will create emergency tables that will require the use of a viral drug.

## 1. Introduction

Wuhan has been based on Coronavirus 2 (SARS – CoV – 2), an infectious pneumonia epidemic that has started rapidly in China and has spread to many countries around the world since December 2019 (The Lancet, 2020). On January 30, 2020, the World Health Organization (WHO) announced that the SARS – CoV – 2 epidemic is a critical and international public health problem. Currently, the disease caused by the SARS – CoV – 2 infection in humans has exceeded the outlook in the severe acute respiratory syndrome (SARS) and the Middle East respiratory syndrome (MERS) outbreak in 2002 (Sun et al., 2020).

To date, the total number of cases in the world was 3,349,786 and the total number of deaths was 238 628 (last update 03/MAY/2020 10: 00 hours, WHO, status report-104) and it is still increasing. Coronaviruses (CoV) are RNA viruses. Until December 2019, the CoV family consisted of six human pathogenic species: SARS-CoV and MERS-CoV. Seventh human pathogenic species was added with SARS-CoV-2 (Hui, 2017). The four “endemic” species (HKU1, OC43, 229E, NL63) are of clinical importance so far, often produce cold symptoms and are responsible for about 10% of seasonal airway diseases not caused by the flu. Besides, SARS-CoV and MERS-CoV, which caused severe airway symptoms and diseases associated with a high mortality rate (10–30%, respectively), were limited to a one-time outbreak and were predominantly of regional significance (Sun et al., 2020).

Real-time reverse transcription-polymerase chain reaction (RT-PCR) diagnostic tests have been rapidly developed on SARS-CoV-2 (Wang et al., 2020). Through genetic sequence analysis, it was stated that SARS-CoV-2 belongs to the genus β-coronavirus with 79.0% nucleotide similarity and 51.8% identity with MERS-CoV and SARS-CoV (Ren et al., 2020). Besides, it has been reported that SARS - CoV - 2 is 96% identical to a bat coronavirus throughout its genome (J. Liu et al., 2020; Zhou et al., 2020).

The pandemic spreading can be fatal when healthcare professionals are not ready to manage the infection, as now seen in the COVID-19 outbreak. The SARS-CoV-2 virus was also isolated from asymptomatic individuals, and affected patients showed contagiousness even 2 weeks after symptoms ceased (Dashraath, Lin, et al., 2020). Thus, it required radical measures on all continents, including closing the country’s borders.

As the epidemic of COVID-19 spreads rapidly, pregnant women have drawn attention to the prevention and control of COVID-19 infection due to being at risk of respiratory infection, especially flu. Physiological and mechanical changes in pregnancy increase susceptibility to infections in general and promote rapid progression to respiratory failure, especially when the cardiovascular system is affected. Thus, they represent a high-risk group during infectious outbreaks (Dashraath, Wong, et al., 2020).

All these risk factors cause an essential point to examine pregnant women. Clinical features and the functionality of the methods of detection of SARS-2 infection in pregnant women are currently the focus of attention in medical studies. Though, owing to the different designs of different clinical trials and small sample sizes, published trials are also various (Sun et al., 2020). The goal of this study was to examine all these articles about the detection of SARS-CoV-2 in pregnant women with molecular (PCR) and CT imaging methods, the frequency of occurrence of these clinical features, and also the detection correction of the methods used in diagnosis by meta-analysis.

## 2. Material and Methods

### 2.1. Sources of Information

MEDLINE PubMed, SCOPUS, Clinical Key Library, CINAHL (Cumulative Index to Nursing and Allied Health Literature) and ISI Web of Science were searched using combined keywords: 2019- nCoV and/or pregnancy”, “COVID-19 and/or pregnancy” and “SARS-CoV-2 and/or pregnancy”.

### 2.2. Article Selection and Publication Quality Evaluation

Meta-analysis was elaborated according to the PRISMA guidelines (Moher et al., 2015). The literature search and selection process are presented in Figure 1, which was conducted according to the PRISMA flow chart.

**Figure 1.**
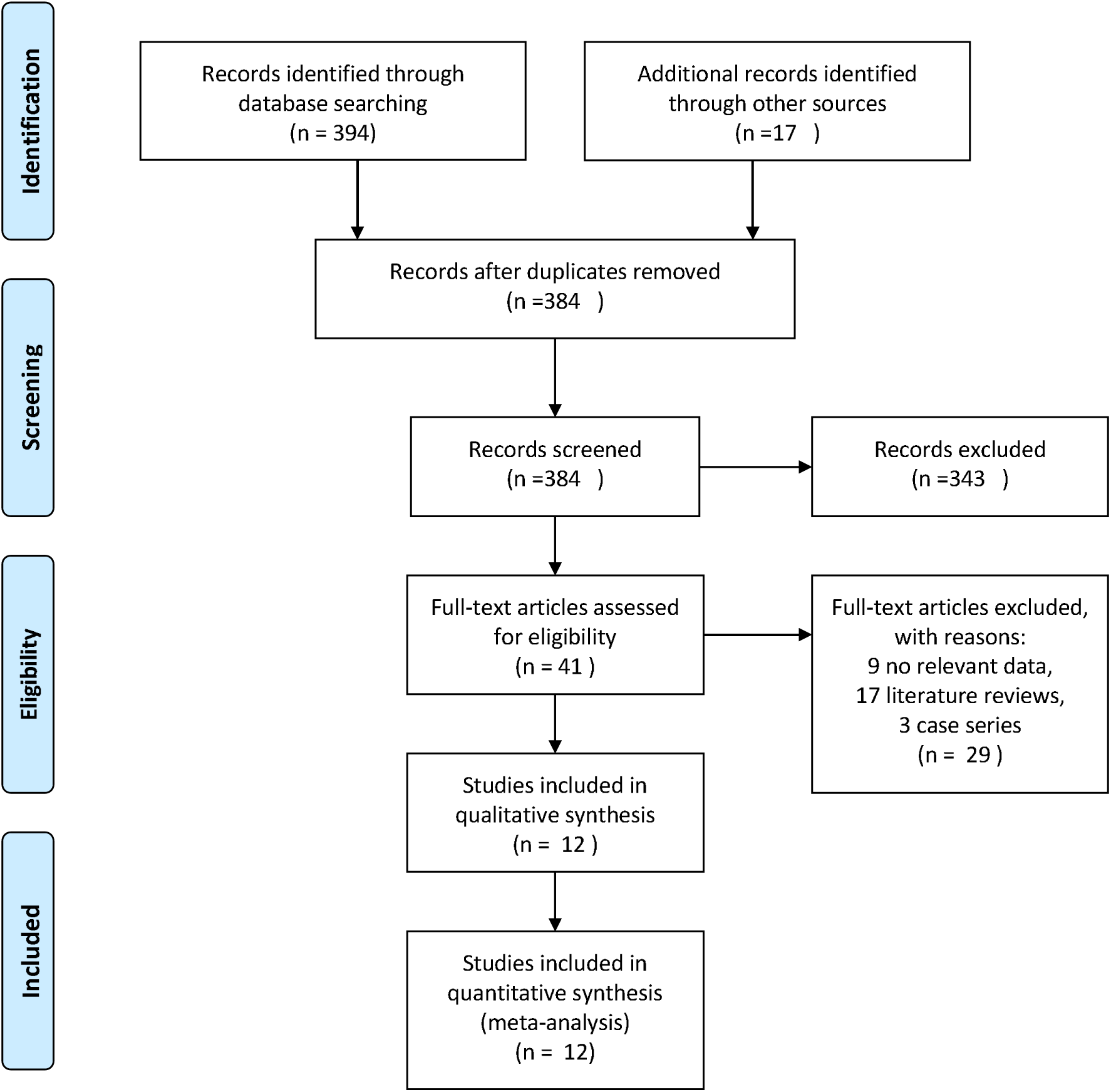
PRISMA flow chart

As a result of the electronic database search, we received 394 articles and 17 of them were excluded due to repeated access. Finally, 12 unique studies were selected that reported on clinical properties and diagnostic methods. The meta-analysis was done using 12 articles (H. Chen et al., 2020; S Chen et al., 2020; Siyu Chen et al., 2020; N. Li et al., n.d.; D. Liu et al., 2020; H. Liu et al., 2020; C. Wu et al., 2020; X. Wu et al., 2020; Yang et al., 2020; Yu et al., 2020; Zhang et al., 2020; Zhu et al., 2020) with a total of 181 patients that satisfied the study criteria. The Newcastle-Ottawa Scale (NOS) was used to assess the quality of included studies (Stang, 2010). The quality scores of all varied from 0 to 9 (Table 1).

**Table 1.**
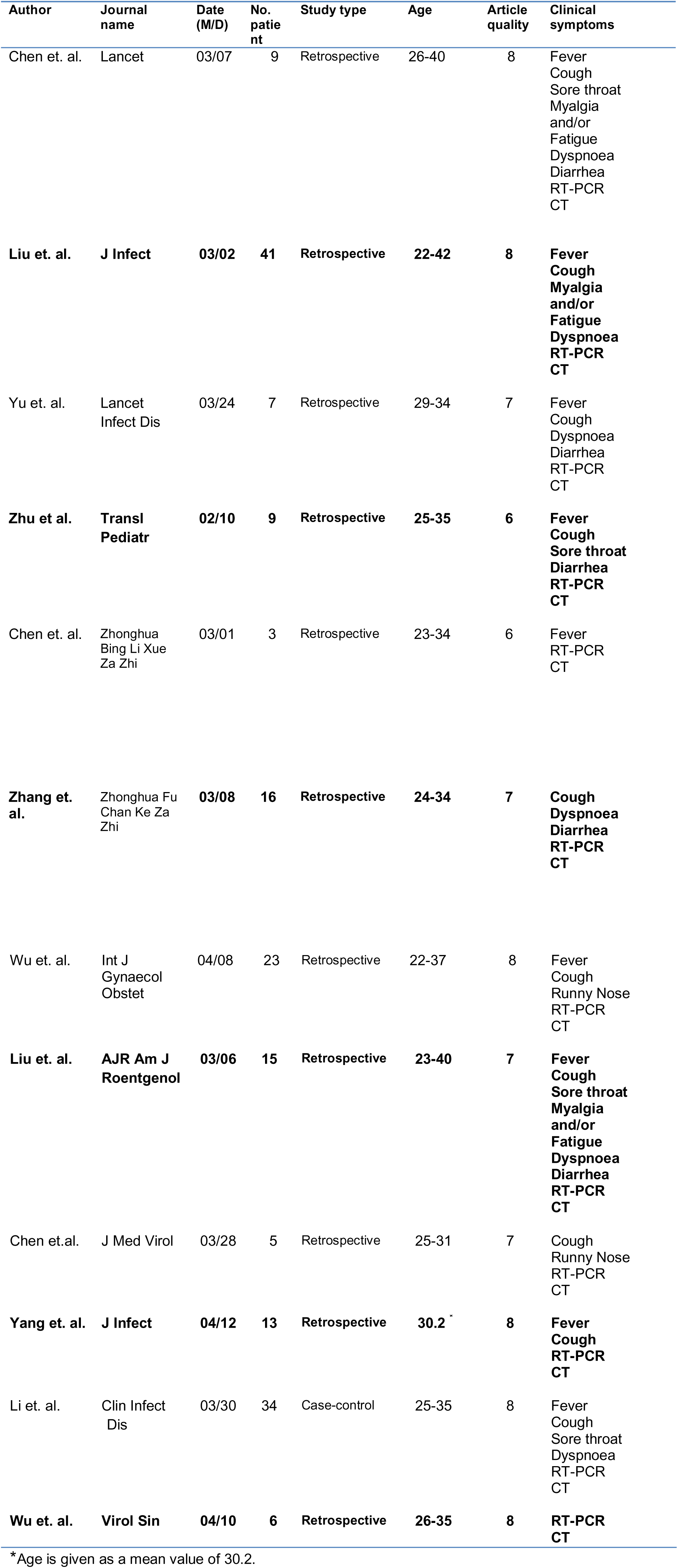
Characteristics of the included studies on COVID-19, 2020.

| Author | Journal name | Date (M/D) | No. patient | Study type | Age | Article quality | Clinical symptoms |
| --- | --- | --- | --- | --- | --- | --- | --- |
| Chen et. al. | Lancet | 03/07 | 9 | Retrospective | 26-40 | 8 | Fever<br>Cough<br>Sore throat<br>Myalgia<br>and/or<br>Fatigue<br>Dyspnoea<br>Diarrhea<br>RT-PCR<br>CT |
| Liu et. al. | J Infect | 03/02 | 41 | Retrospective | 22-42 | 8 | Fever<br>Cough<br>Myalgia<br>and/or<br>Fatigue<br>Dyspnoea<br>RT-PCR<br>CT |
| Yu et. al. | Lancet Infect Dis | 03/24 | 7 | Retrospective | 29-34 | 7 | Fever<br>Cough<br>Dyspnoea<br>Diarrhea<br>RT-PCR<br>CT |
| Zhu et al. | Transl Pediatr | 02/10 | 9 | Retrospective | 25-35 | 6 | Fever<br>Cough<br>Sore throat<br>Diarrhea<br>RT-PCR<br>CT |
| Chen et. al. | Zhonghua Bing Li Xue Za Zhi | 03/01 | 3 | Retrospective | 23-34 | 6 | Fever<br>RT-PCR<br>CT |
| Zhang et. al. | Zhonghua Fu Chan Ke Za Zhi | 03/08 | 16 | Retrospective | 24-34 | 7 | Cough<br>Dyspnoea<br>Diarrhea<br>RT-PCR<br>CT |
| Wu et. al. | Int J<br>Gynaecol<br>Obstet | 04/08 | 23 | Retrospective | 22-37 | 8 | Fever<br>Cough<br>Runny Nose<br>RT-PCR<br>CT |
| Liu et. al. | AJR Am J<br>Roentgenol | 03/06 | 15 | Retrospective | 23-40 | 7 | **Fever**<br>**Cough**<br>**Sore throat**<br>**Myalgia**<br>**and/or**<br>**Fatigue**<br>**Dyspnoea**<br>**Diarrhea**<br>**RT-PCR**<br>**CT** |
| Chen et.al. | J Med Virol | 03/28 | 5 | Retrospective | 25-31 | 7 | Cough<br>Runny Nose<br>RT-PCR<br>CT |
| Yang et. al. | J Infect | 04/12 | 13 | Retrospective | 30.2 * | 8 | **Fever**<br>**Cough**<br>**RT-PCR**<br>**CT** |
| Li et. al. | Clin Infect<br>Dis | 03/30 | 34 | Case-control | 25-35 | 8 | Fever<br>Cough<br>Sore throat<br>Dyspnoea<br>RT-PCR<br>CT |
| Wu et. al. | Virol Sin | 04/10 | 6 | Retrospective | 26-35 | 8 | **RT-PCR**<br>**CT** |

\*Age is given as a mean value of 30.2.

### 2.3. Statistical analysis

The meta-analysis of incidence rates was conducted using the “metafor” package in R version 3.6.2 (Viechtbauer, 2010). It includes 12 studies, with a total of 181 patients. We employed the random-effects model according to assessing heterogeneity of the meta-analysis.

The publication bias was detected by Egger’s test. The test results and corresponding P values are presented in Table 2. The Egger’s test indicated that publication bias exists for diarrhea and RT-PCR groups (P = .019 and P=0.025, respectively)

**Table 2.**
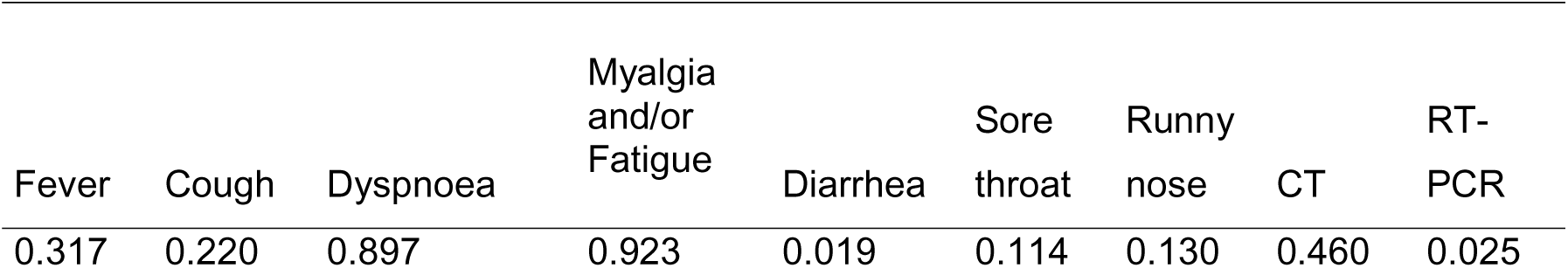
Results of the Egger’s test.

The double-arcsine method would be a more appropriate choice when the small sample size and extreme proportions need to be handled. The double arcsin transformation was applied in order to make the skewed distribution of proportions to conform to the normal distribution (Barendregt et al., 2013; Rousseau & Evans, 2017). To conclude, we performed the inverse of the double arcsine transformation for proportions using the harmonic mean of the sample sizes for the back-transformation. The results are given for the summary incidence rate and confidence interval in Table 3.

## 3. Results

The incidence of fever was 38% (95% CI: 14% –35%) and cough was 22% (95% CI: 10.8 –35.2%) among all clinical features of pregnant cases with SARS-CoV-2 infection by meta-analysis. Dyspnoea was observed only 3.3% (95% CI: 0.3 –8.2%). The incidence of positive RT-PCR is 91.8 % (95% CI: 76.7–99.9%) and the incidence of abnormal computer tomography (CT) is 97.9% (95% CI: 94.2–99.9%).

Since no clinical signs of vomiting were found in any of the studies included in the analysis, the effect size was not given for it.

Detailed results of the meta-analysis are shown in Table 3.

**Table 3.** Meta-analyses results.

| Clinical symptom | Results of<br>Meta-analysis | Lower limit | Upper limit |
| --- | --- | --- | --- |
| Fever | 0.381 | 0.142 | 0.650 |
| Cough | 0.220 | 0.108 | 0.352 |
| Dyspnoea | 0.033 | 0.003 | 0.082 |
| Myalgia and/or<br>Fatigue | 0.030 | 0.000 | 0.115 |
| Diarrhea | 0.004 | 0.000 | 0.036 |
| Sore throat | 0.002 | 0.000 | 0.028 |
| Runny nose | 0.000 | 0.000 | 0.010 |
| CT | 0.979 | 0.942 | 0.999 |
| RT-PCR | 0.918 | 0.767 | 0.999 |

## 4. Discussion

In the early stages of the SARS-CoV-2 epidemic, the case death rate is estimated to be approximately 2% (Sun et al., 2020). Later, it is reported that the death rate in China was 3.8%, which was lower than that of the two commonly transmitted zoonotic CoV diseases, SARS and MERS (J. Liu et al., 2020).

SARS-CoV-2 infection is more likely to affect older people with comorbidity, with most deaths clustering in this particular population (N. Chen et al., 2020; Q. Li et al., 2020; J. Liu et al., 2020). The mortality rates of SARS and MERS infections are 9.6% (Hui, 2017) and 35% respectively (Azhar et al., 2019). Xie et al (Xie et al., 2005), stated that 45% of patients showed symptoms of pulmonary fibrosis within 1 month after infection with SARS-CoV, and 30–36% after 3–6 months. These studies suggested that pulmonary fibrosis will be one of the serious complications in patients with SARS-CoV-2 infection. Furthermore, due to the low mortality rate of SARS-CoV-2 infection and rapid spread among patients compared to SARS-CoV and MERS-CoV infections, a large number of patients require treatment. In this case, health equipment and health worker competence has been essential. Thus, COVID-19 threatens preparedness and biosecurity conditions in all countries. (Rodriguez-Morales et al., 2020). At the same time, both of these coronaviruses can cause death in a few, but significant numbers of pregnant cases, but specific risk factors for a fatal outcome during pregnancy have not been clarified (Sun et al., 2020).

In addition, in another study, Zhao et al. (2020), the angiotensin-converting enzyme 2 (ACE2) SARS-221-CoV-2 receptor, they found that in healthy lung tissue, ACE2 is mainly expressed by type I and type II alveolar epithelial cells. Type II alveolar cells (83%) have been reported to express ACE2. For this reason, infection damages most of the type II alveolar cells. The use of mechanical ventilation in the treatment of patients can also aggravate the damage of alveolar cells. However, it has also been reported that ACE2 is more expressive in pregnant women (Pringle et al., 2011; X. Zhao et al., 2020).

Compared with past coronavirus pandemics, it has been reported that pregnancy has a significant impact on the course of the disease of SARS-CoV and the outcome of an infected patient. Therefore, the duration of the hospital stay of pregnant patients was longer. In addition to kidney failure, sepsis, or common intravascular coagulation disorder, the need for intensive care treatment was more common in pregnant women. The mortality of pregnant infected patients has also increased significantly (Lam et al., 2004). So far, there is very little data on MERS-CoV infection during pregnancy. However, 11 reported symptomatic cases (Alfaraj et al., 2019), showed a more severe course in pregnancy than SARS-CoV infection.

In addition to information about the effects of previous coronavirus outbreaks on pregnant women, little data is known about the clinical course, possible risks, and the validity of the methods used in the correct diagnosis for pregnant patients infected with SARS-CoV-2. In this study, a meta-analysis of the data of publications examining these possible risks and methods used in diagnosis for pregnant women suffering from SARS-CoV-2 infection was performed. The results we found in pregnant women with SARS-CoV-2 infections are common symptoms of fever, cough, shortness of breath, general myalgia, weakness, diarrhea, dyspnea, and pneumonia compared to the primary clinical symptoms in pregnant women with MERS-CoV and SARS-CoV infections (J. Liu et al., 2020).

When we compare our results of Meta-analysis for pregnant women with the rates of the study conducted with the non-pregnant adult group (Sun et al., 2020), respectively, the incidence of fever was 38% (89.1%), the cough incidence 22% (72.2%) and the incidence of myalgia and/or fatigue 3% (42.5%). The incidence of dyspnoea is 3.3% (14.8%), the incidence of abnormal CT is 98% (96.6%), and the case fatality rate of patients with SARS-CoV-2 infection is 4.3% (H. Chen et al., 2020; S Chen et al., 2020; Siyu Chen et al., 2020; N. Li et al., 2020; D. Liu et al., 2020; H. Liu et al., 2020; C. Wu et al., 2020; X. Wu et al., 2020; Yang et al., 2020; Yu et al., 2020; Zhang et al., 2020; Zhu et al., 2020). In addition, symptoms of diarrhea, sore throat, and runny nose are rare. In no case, mechanical ventilation was not required in pregnant women, also there were no reported cases of death.

All irregularities in the imaging results were considered abnormal for the CT result. According to our RT-PCR and CT results (91.8% –97.9% respectively), we think that both should be used when evaluating to say that the cases are correct and definite positive in pregnant women. However, it should be noted that there is radiation in the CT examination, so the question of whether the re-examination interval is necessary in the treatment of pregnant women with mild symptoms needs further discussion. Based on the above, COVID-19 has mild clinical signs in pregnant women; some are asymptomatic and need to be combined with epidemiological history and nucleic acid detection. Weijie Guan et al. (2020), stated that frequent patterns in chest CT include abnormal findings in the case reports, including asymptomatic patients, with ground-glass opacity and bilateral irregular shading (Chan et al., 2020; Huang et al., 2020; J. Liu et al., 2020). Weijie Guan et al. (2020), stated that frequent patterns in chest CT include abnormal findings in the case reports, including asymptomatic patients, with ground-glass opacity and bilateral irregular shading (Chan et al., 2020; Huang et al., 2020; J. Liu et al., 2020).

The results of this meta-analysis study here highlight the clinical, molecular, and imaging findings of COVID-19 pregnant cases that may assist clinicians. In this way, it will prevent further contamination by implementing infection control measures, thanks to early recognition of cases and adequate intervention by clinicians. Frequent evaluation of available evidence of COVID-19, such as clinical suspicion and definitive diagnosis, has been deemed necessary to prevent contamination from health workers during close contact in pregnant women(Biscayart et al., 2020; Holshue et al., 2020; Rodríguez-Morales et al., 2020).

Here we have discussed 12 articles, including 181 pregnant cases with SARS-CoV-2 infection. So far, it is the first meta-analysis to examine the factors and prenatal clinical features that may be effective in initial diagnosis in pregnant women. The quality of the literature included in this study is high. However, this review also has some limitations. As of the current period, there are few studies for the content. Data from all countries are urgently needed on this issue. Thus, it would be more appropriate to include a large number of studies in a broad geographical scope in order to obtain a more comprehensive view of COVID-19 in pregnant women as a result. Since detailed patient information was not given in all studies, especially regarding clinical findings, this could not be included in the meta-analysis. In particular, there were negative results, although the case showed positive clinical signs, since CT contained radiation, and its use was not preferred or repeated. The data in this analysis allow for the first synthesis of the clinical, molecular, and CT diagnostic features of COVID-19. Also, it is not included in the meta-analysis results, since deaths were not reported in pregnant women in the studies conducted. In this study, the patients were diagnosed with SARS-CoV-2 infection. Therefore, asymptomatic cases are not emphasized. However, because the clinical symptoms are rare in the findings of our study, the prevalence may be higher among pregnant women. As there is a lack of data in newborns as part of the studies we have included, it could not be covered in this study. As a result, the importance of vertical transmission is not emphasized.

Based on the limitations reported above, the results need to be supported by more extensive studies with larger sample sizes. Further clinical data are essential to explain the clinical spectrum of the disease. Clinical experience case reports, case series or large observational studies from countries with an increasing number of cases will contribute significantly.

Pregnant women are sensitive to respiratory pathogens and the development of severe pneumonia, making them more susceptible to COVID-19 infection, especially if they have chronic diseases or complications (Qiao, 2020). Therefore, pregnant women should be considered as an critical risk group in the prevention and treatment of COVID-19 infection. In addition to recent studies, previous pandemic experiences should be considered in the prevention and control of this infection. Our findings will provide valuable guidance for current clinical trials. We are also of the opinion that our results can give an idea about the necessity of using a viral drug in the treatment of SARS – CoV – 2 infections in pregnant women. In summary, pregnant women with SARS-CoV-2 infection should be closely monitored for early diagnosis. Currently, pregnancy may complicate the clinical course of COVID-19, but the fact that the cases in this group are not in the risky age group defined for COVID-19 may give an idea that their condition will not be as bad as in the pregnant tables in MERS or SARS infections (Sun et al., 2020).

## 5. Conclusion

We believe that this research may be critical in determining methods and even saving lives in the early diagnosis and treatment of pregnant women in current and future outbreaks.

## Data Availability

The data that support the findings of this study are openly available in figshare at https://doi.org/10.6084/m9.figshare.12442031

## Data Availability

All data generated or analyzed during this study are included in this article.

## Authors’ Contributions

BUC designed the research model and determination of these articles to be included in the study. BAB contributed to the realization of the study by applying meta-analysis. Both authors contributed to article selection and data interpretation, reviewed and approved the final article.

## Acknowledgments

This work was not supported.

We would like to express our gratitude to all healthcare professionals working hard on these challenging days.

## Ethical Approval

Approval was not required.

